# PROJECTING A MATURE EPIDEMIC: A SIMPLE TOOL WITH AN APPLICATION TO COVID-19 DEATHS

**DOI:** 10.1101/2020.07.13.20152512

**Authors:** Marc Artzrouni, Randy Wykoff

**Author notes:** **Conflict of interest:** none. **Data/code:** data from usafacts/org; VBA code available in Excel sheet provided in Supplementary Material.

## Abstract

We describe a new statistical model for the spread of a mature epidemic, i.e. one characterized by an exponentially decaying growth rate of the cumulative number of cases/deaths – the speed of this decay being measured by the growth rate’s half-life. If such a pattern is observed during the recent past, then it can be extrapolated. A spreadsheet is made available that allows users to input weekly cumulative numbers of deaths and obtain an estimate of the growth rate’s baseline half-life and the corresponding projections. These projections can be compared to those with a larger half-life (if a protracted epidemic is expected, e.g. due to second wave), or with a smaller one (if successful therapies or mitigation efforts reduce transmission). The model is applied to deaths due to COVID-19 in California in May-June 2020.

---

A recent note^1^ in this journal describes a Bayesian model to estimate numbers infected in the early exponential phase of the COVID-19 outbreak (the growth rate of total cases is then constant). Here we use a new statistical model^2^ to project the future course of a “mature epidemic”, i.e. one whose growth rate (of either total cases or deaths) is no longer constant but decays exponentially. Many European countries have followed this pattern during the COVID-19 pandemic, with a mature period well underway by April or May 2020. This followed an early exponential phase that could be surprisingly short.

The earliest affected U.S. states (California, Washington State and several states in the northeast) experienced a similar pattern at about the same time. Other states were affected later and differently as the pandemic spread outside large metropolitan areas such as New York City.

We proceed with a question that could have been asked by the California Department of Public Health (CDPH) on May 21: “As of today there have been 3,597 COVID-related deaths in the state, a 17.97% increase over the previous week’s total of 3,049. Is there a way we can predict an ultimate number of deaths during this outbreak?” The formulas given below answer this question under the mature epidemic assumption.

## METHOD

The new model mentioned above is called the “Exo-r” model – where this terminology refers to a growth rate *r(t)* that changes exogenously with time *t*. Specifically, *r(t)* is constant during the early exponential phase and decreases exponentially thereafter, during the mature phase, as discussed above. The model was successfully applied to *total cases* for the completed COVID-19 epidemic in China (January to April 2020) – and to numbers of *reported deaths* in the U.S. up to May 2020^2^.

Here we will focus on the cumulative number of deaths *D(t)* in an ongoing, mature COVID-19 epidemic. We thus consider an initial “time of maturity” *t*=0 from which *r(t)* decays exponentially at some (negative) rate *s*, i.e. *r*(*t*) = *r*_0_*e*^*st*^. Under these assumptions the total number of deaths up to time *t*>0 is

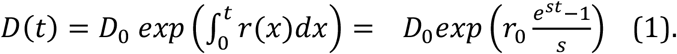

We choose a weekly time step which smooths daily numbers of COVID-19 deaths. The initial (instantaneous) growth rate *r*_0_ is approximated by ln 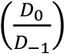 where *D*_−1_ are total deaths up to week -1. The (initial) weekly growth rate calculated by CDPH is 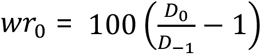.

The rate of decay of the growth rate can be measured by its half-life *HL*, i.e. the time it takes for it to halve. The half-life is easier to interpret than the growth rate *s* and is equal to HL=−ln(2)/*s* (remember s is negative). Under the effect of an exponentially decaying growth rate, deaths *D(t)* of Eq. (1) increase more and more slowly with time, reaching for t→∞ the limiting value

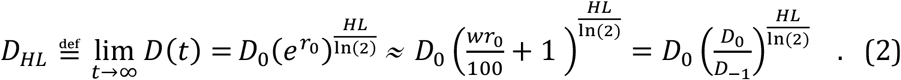

In our example *D*_0_ = 3,597 and w*r*_0_ = 17.97% : the only thing CDPH needs in order to project deaths is the half-life *HL* - hence the *D*_*HL*_ notation. A small *HL* means a growth rate that will rapidly decay to zero. The exponent is then close to 0 and the epidemic will come to a swift end since *D*_*HL*_ is close to *D*_0_. A large *HL* means a growth rate that decays slowly, resulting in a protracted epidemic and a large toll *D*_*HL*_.

Given that *r*(*t*) = *r*_0_*e*^*st*^ for *t*>0, we have ln(*r*(*t*)) = ln(*r*_0_) + *st*. The decay rate *s* prevailing in the recent past is obtained as the estimate *ŝ* of the slope in the simple linear regression of weeks vs ln(*r*(*t*)) over a few weeks prior to time 0 (Figure 1). As before each growth rate *r(t)* (*t* =−1,−2,…) is estimated as ln 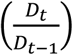. The estimated half-life is 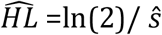. Extrapolating *r(t)* beyond *t*=0 with the decay rate *ŝ* yields the projected total deaths *D(t)* of Eq. (1).

**Figure 1.**
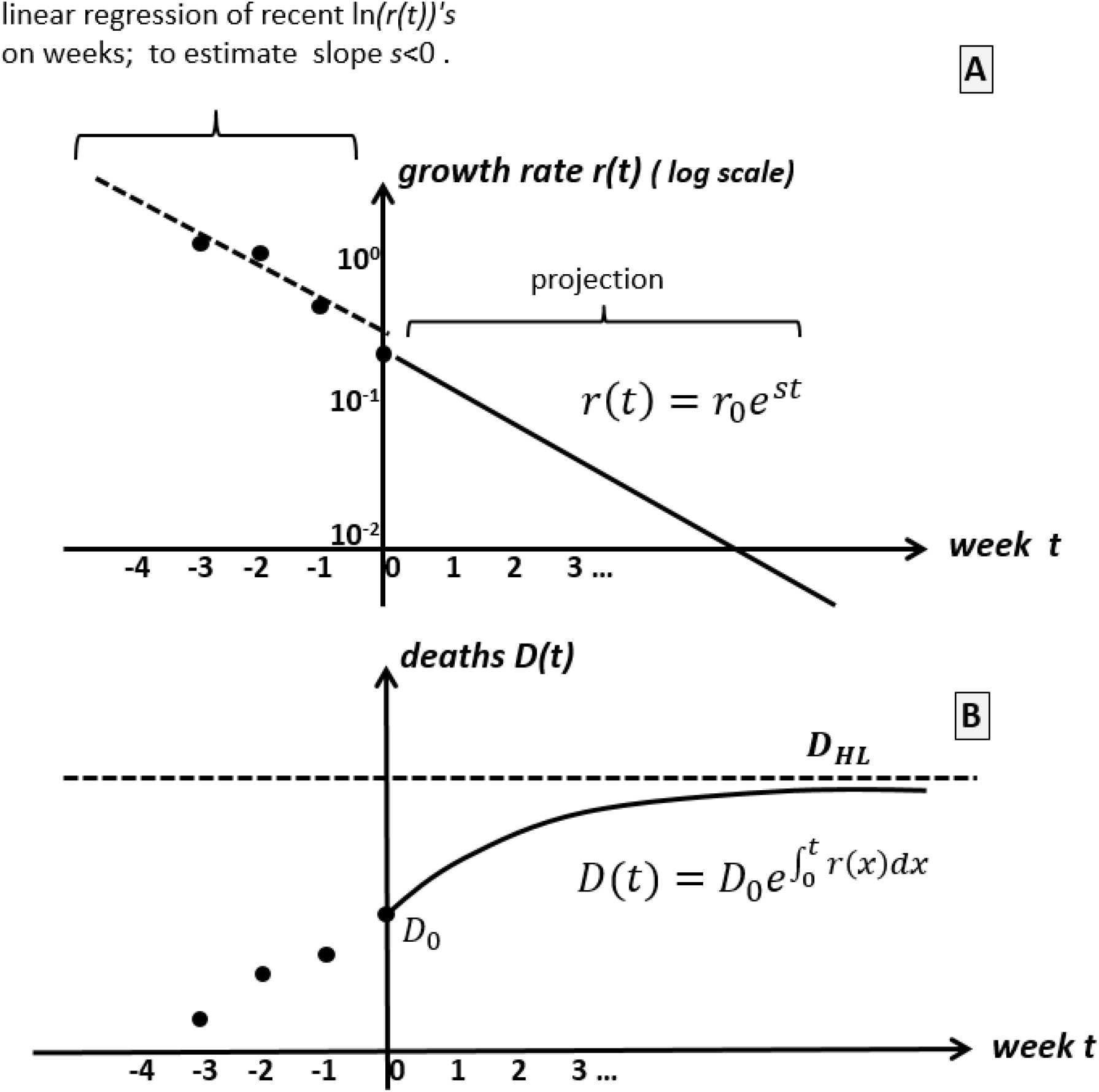
Projection of a mature epidemic in which the growth rate *r(t)* of total deaths *D(t)* decreases exponentially (Panel A). The linear trend prior to week 0 is extrapolated, resulting in cumulative numbers of cases *D(t)* that increase to the limit *D*_*HL*_ (Panel B).

## APPLICATION

We first assume that today is May 21(*t*=0) and CDPH wishes to project the numbers of deaths beyond that date. The linear regression performed over the five-week period (six data points) from April 16 to May 21 yields the baseline 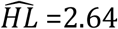 weeks with a coefficient of correlation *R*= −0.98 (Figure 2). Extrapolating the number of deaths with this value of *HL* leads to an ultimate number of deaths *D*_*HL*_ = 6,751. (See Exo-rProjection Excel workbook available from Supplemental Digital Content 1).

**Figure 2.**
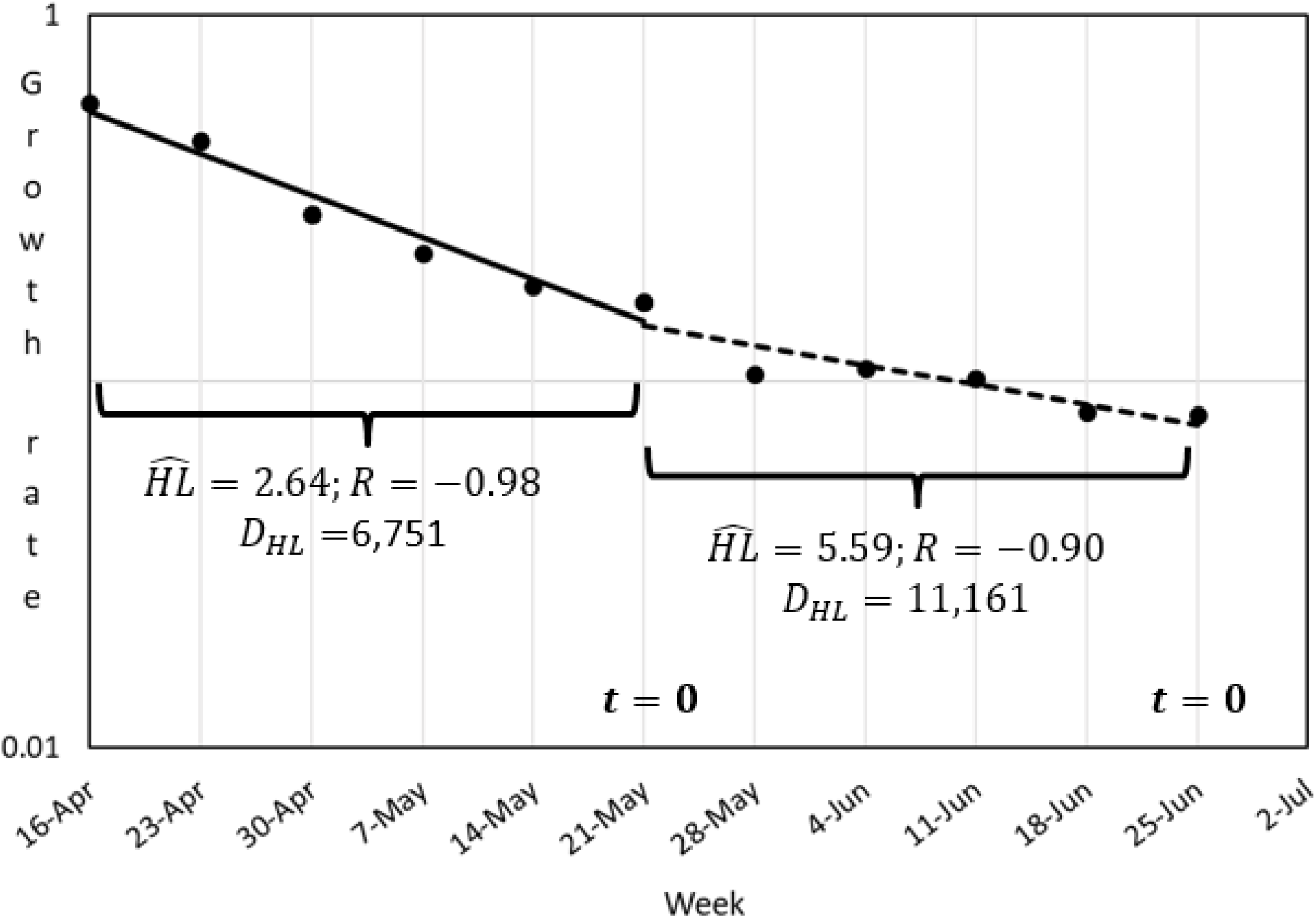
Linear regression of weeks vs growth rate (logarithmic scale) of deaths in California performed separately for two six-week periods: i) April 16 to May 21 (*t*=0); and ii) May 21 to June 25 (updated *t*=0). The longer estimated half-life *HL* during the second period reflects a growth rate that was decreasing more slowly than during the previous period. The projected outbreak is thus more protracted than was predicted on May 21, with an ultimate number of deaths *D*_*HL*_ that almost doubles.

Fast forward to June 25 now taken as time *t*=0. (Figure 2). By then the number of deaths in California had reached 5,804. This suggests that the ultimate total of 6,751 total was perhaps optimistic. We performed the same linear regression as above, over the five-week period from May 21 to June 25. Although the line appears only slightly less steep from May 21 to June 25, the half-life has more than doubled to 5.59 weeks. Extrapolating this value beyond June 25 means that the growth rate will decay more slowly than before resulting in a larger *D*_*HL*_ of 11, 161 calculated from Eq. (2) (Figure 3). This is more than twice the previous ultimate number of 6,751.

**Figure 3.**
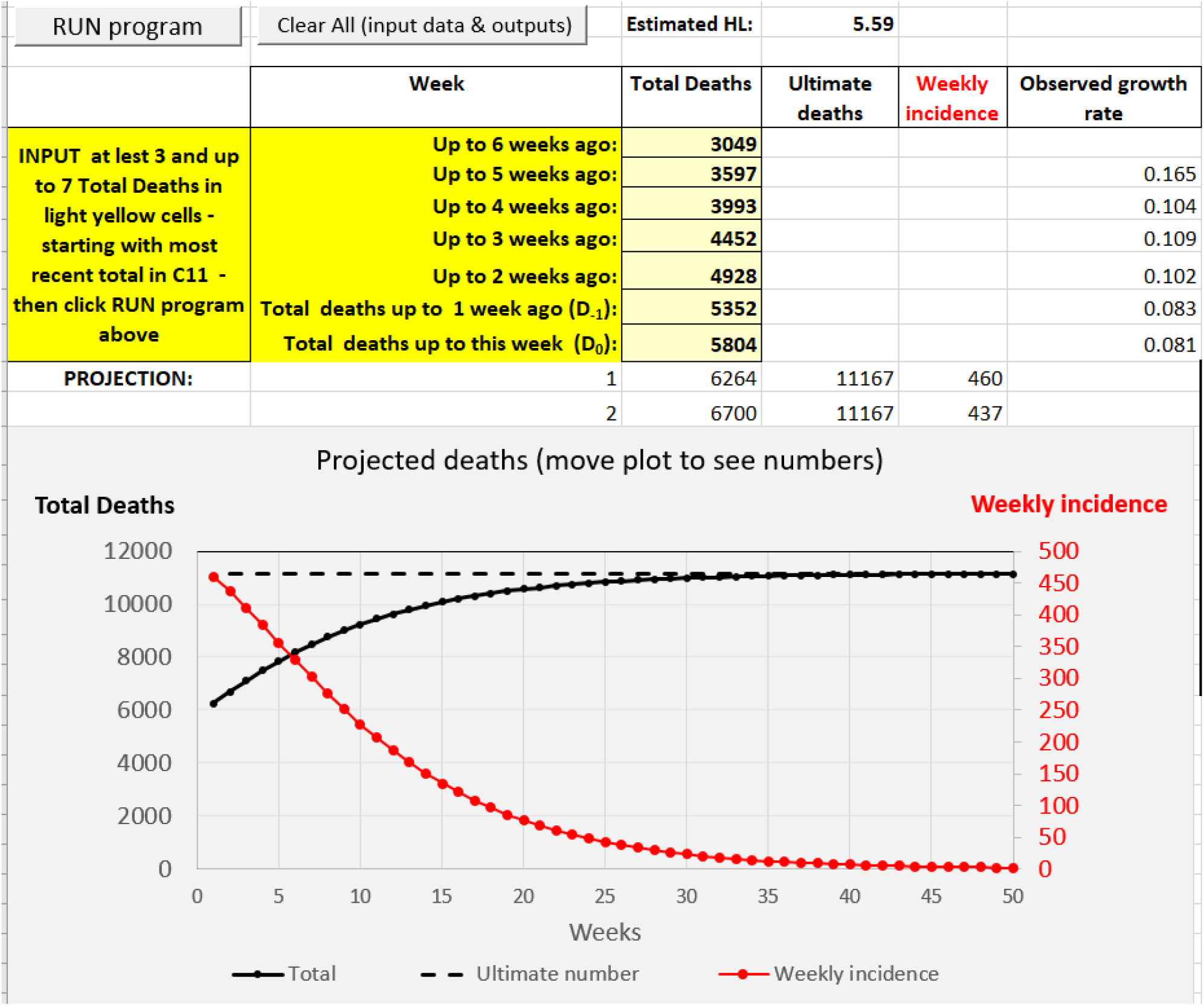
Partial view of output obtained with the Exo-rProjection Excel sheet for deaths in California from June 25, 2020 (Week *t*=0) at which time the total number was *D*_*0*_=5,804. The growth rate decays with an estimated half-life of 5.59 weeks. Total cases approach the limit *D*_*HL*_ = 11,167.

The projection beyond June 25 does not have to assume the recently prevailing 5.59 weeks half-life. The implications of a growth rate that may decrease more slowly (e.g. because of a resurgence of cases) can be explored by assuming values of *HL* larger than 5.59. Conversely, therapeutic improvements could accelerate the decrease meaning a shorter *HL*.

## DISCUSSION

Any projection is a “what-if” exercise – the “what-if” here being a continued exponential decay in the growth rate following the exponential phase. Our projections for California illustrate the method’s simplicity as well as its limits: results are sensitive to the assumed half-life – which may also have to be updated. But this is easily done with a simple interactive spreadsheet that requires only a handful of weekly numbers of deaths.

It will be challenging to apply the method described here to COVID-19 cases in the U.S. given the severe under-reporting of these cases and the vagaries of testing protocols. Still, *if* reported cases *consistently* represent only 10% of all infected people, then the projection could arguably reflect 10% of future cases. In July 2020 U.S. cases and deaths are moving in opposite directions (unlike in Europe): the continued expansion of the epidemic brought about by the premature reopening of the economy is causing cases to surge in many states while improved therapies are driving the number of deaths down. The epidemic of cases in the U.S. may therefore not be mature enough to be projected with the model described here – but could be in the future.

## Data Availability

data and program available

## Supplementary Digital Content 1

Has explanations and link to “Exo-rProjection” Excel workbook (https://bit.ly/3bAYFZr). There are two sheets. One uses the linear regression described above to calculate the (baseline) doubling time *HL* and project deaths accordingly. The other requires only two weeks of data and a proposed half-life *HL*. This allows for the exploration of diverse scenarios concerning the rate of decay of the growth rate.

## Notes

**Financial support:** none.

### Competing Interest Statement

The authors have declared no competing interest.

